# Implicit, Intrinsic, Extrinsic (or Environmental), and Host Factors Attributing the Covid-19 Pandemic. Part 2-Implicit Factor Pesticide Use: A Systematic Analysis

**DOI:** 10.1101/2021.09.09.21263347

**Authors:** Ananya Aggarwal, Ragini Rai, Gaurav Joshi, Prashant Gahtori

## Abstract

Advances in our understanding of complex COVID-19 pandemic would allow us to effectively eliminate and eradicate SARS-COV2 virus. Although tremendous amount of research devoted to the robustness across its biology, diagnostics, vaccines and treatment has exploded in the past two years. However, still science do not have robust answers for causes, for example (i) What are the reasons of non-uniform global distribution of COVID-19? (ii) Why the United States, India, and Brazil, are the first-three most affected nations?, (iii) How did Bhutan, a nation sharing a boundary with China manage nearly 0.34% infections and 3 deaths from COVID-19? Nonetheless, the biomass bistribution of biosphere report suggest more than 1550-fold larger microbial biomass involving bacteria, fungi, archaea, protists and viruses is exist in comparision to all global human population in the biosphere. The rich microbiota act a first line of defence to invade pathogens and affect us both through the environment and microbiome. Unfortunately, a role of pathogen-transmission factors *viz*. implicit factors (competitive microflora) is still under represented. This study is an attempt from a gold standard correlation methodology using a large pesticide use global data. The non-specific pesticides kill both pests as well as protective microbiota, resulting a loss in rich biodiversity and allow easy pathogen entry to human. Entire predictions were found consistent with the recently observed evidences. These insights enhanced scientific ability to interrogate viral epidemiology and recommended to limit pesticide use for future pandemic prevention.

## Introduction

The origin of life on earth is a natural process ranging from non-living microorganisms (viruses and viroids) to living micro-/macro-organisms (bacteria, archaea, fungi, protists, plants, animals and humans). The microorganisms are found almost in every habitat present in nature and their coexistence with the environment determines the health of the entire ecological system of earth [1]. Soil is an important habitat for both bacteria and cyanobacteria due to an abundance of organic carbon on earth. The robustness of prokaryotic life in the deeper soil subsurface below 8 m, is also enormous (10^3^–10^6^ cells/cm3), and biomass of prokaryotes are detected even at higher altitudes 57–77 km. Airborne prokaryotes also represent a large fraction with 5 × 10^19^ cfu[2]. The average microbial density on earth is approximately 10^8^ microbes/ml, which showed inseparable interactions between humans and microbes everywhere on the planet. Particularly, the human itself contains trillions microbiome in a surprising ratio of 10:1 with human cells. A huge number of a total of 10^30^ prokaryotes exist on earth, out of about 1400 (negligible 1.4×10-25%) microbial species are recognized as human pathogens[3]. Interestingly, a report of the biomass distribution of large Biosphere suggests though large, human population 0.06 gigatons of carbon (Gt C), is still negligible in comparison to bacteria 70 Gt C, fungi 12 Gt C, archaea 7 Gt C, protists 4 Gt C and viruses 0.2 Gt C [4]. Hence, a large number of ecological communities of microorganisms, so-called microbiota with a high level of adaptation and evolution, exist in the environment which protects humans from pathogenic microorganisms and toxins. Unfortunately, human activities over the years via antibiotics, agri-/industrial chemicals, and lifestyle have significantly altered biodiversity[5]. In fact, several times warnings to humanity from leading global scientists including Nobel laureates are well documented[6-9]. The scientific report also highlighted 58% of the earth’s land coverage already falls below safe level[10] and merely ten months old UN report stipulates up to 1million species are at risk of annihilation[11]. Thus, a well-balanced ecosystem products/services usually reduce pathogen transmission, and several studies suggest new virus infections may be a result of loss in biodiversity[12-14].

The modelling of infectious disease dynamics remains the ideal research design and it is essential to identify the key factors responsible for any communicable disease transmission in population level. The epidemiological model usually improves the scientific ability by two-fold: (i) to interrogate viral epidemiology, and (ii) to run counter operations for moving forward all with more accuracy, precision, and reliability. Previous studies also suggest the importance of four crucial transmission factors for any microbial infections, such as (i) intrinsic factors (related to direct food supply to microorganism, for example, nutrient content); (ii) extrinsic or environment factors (external to the food, for example, temperature); (iii) implicit factors (competitive microflora); and (iv) host factors (for the spread of disease, for example, malnutrition)[15,16]. The rising population has significantly impacted the human activities, and we assume fertilizer consumption/nitrous oxide emissions-which itself supplies nutrients, and non-specific pesticides-itself inhibits microflora, could be a part of intrinsic-and implicit-factors respectively. The COVID-19 disease caused by novel corona (SARS-COV2) virus has posed serious health challenges with enormous morbidity and mortality across the world. Till date of writing this report COVID-19 has claimed a total of 185.29153 million confirmed cases and the mortality is again on constant surge with each passing day. With enough scientific evidences reported till date top ten COVID-19 affected countries with cumulative total cases, (i) United States of America (33451965), (ii) India (30752950), (iii) Brazil (18909037), (iv) Russian Federation (5733218), (v) France (5686066), (vi) Turkey (5465094), (vii) The United Kingdom (5022897), (viii) Argentina (4593763), (ix) Colombia (4426811) and (x) Italy (4267105) [17]. The knowledge gap insists us to explore role of key factors over COVID-19 progression as an ideal model system. Hence, a wider sampling and subsequent data analysis of current COVID-19 infectious cases with Pesticides including Fungicides and Bactericides, Insecticides, and Rodenticides use countrywise were envisaged in understanding the epidemiology of COVID-19 and create an unprecedented opportunity for COVID-19 prevention in restoring good quality of life.

## Methods

All COVID-19 infectious cases were collected from five WHO situation reports-dated from 11-Jul-2021 to 28-Mar-2020[17]. The data pertaining to pesticide use countries/ territories-wise was free to use and supplied by the Food and Agriculture Organization (FAO) [18]. Here, the Countries/ Territories with missing entries were omitted, and 98 countries randomly fitted to top variable indices of correlation coefficient for unbiased cause-effect relationships.

## Results and Discussion

There is considerable evidence that intrinsic, extrinsic (or environment), implicit, and host factors are key regulators for pathogen transmissions. In this study, we have observed a total pesticide use was optimally correlated with the number of COVID-19 infected cases, as appeared on March 13th (r^2^=0.93), 19th (r^2^=0.86), and 28th (r^2^=0.57), 2020 (Table 1). Later on consistant correlation (0.51≤r^2^ value≤ 0.54) was seen in case of a total fungicides and bactericides from last one year i.e. March 28th, 2020 to July 09th, 2021 with a very weak standard error value 0.02. From December 22th, 2020 a high correlation was witnessed in case of a total insecticides use with r^2^ value ranges from 0.80 to 0.83. The United States of America, Brazil, Thailand, and India are among top four insecticides consumer countries and currently United States of America, India, and Brazil are top three COVID-19 affected nations. Other hand, as per FAO report Bhutan do not utilize insecticides and Brunei Darussalam, consumed very limited insecticides, a total of 1 and 3 tonnes only. Both Bhutan, and Brunei Darussalam are among top two least COVID-19 affected nations. These real evidences are advised the direct role of pesticides use on increasing COVID-19 infected cases.

**Table 1.** Cumulative COVID-19 Infected Cases and Pesticides Consumption Worldwide

|  | Country/<br>Territory/Area | CUMULATIVE COVID-19 INFECTED CASES |  |  |  |  | Total<br>Pesticides<br>(Tonnes) | Total<br>Fungicides<br>and<br>Bactericides<br>(Tonnes) | Total<br>Insecticides<br>(Tonnes) | Total<br>Rodenticides<br>(Tonnes) |
| --- | --- | --- | --- | --- | --- | --- | --- | --- | --- | --- |
|  |  | 09-Jul-21 | 22-Dec-20 | 13-Mar-20 | 19-Mar-20 | 28-Mar-20 |  |  |  |  |
| 1 | Albania | 132565 | 52542 | 23 | 59 | 197 | 442 | 199 | 119 | 36 |
| 2 | Algeria | 143652 | 94781 | 25 | 72 | 367 | 6067 | 1577 | 367 |  |
| 3 | Argentina | 4593763 | 1531374 | 31 | 79 | 589 | 172928 | 3427 | 3747 | 0 |
| 4 | Armenia | 226135 | 153825 | 1 | 84 | 372 | 581 | 302 | 143 | 17 |
| 5 | Australia | 30905 | 28128 | 140 | 510 | 3635 | 63416 | 4544 | 14196 |  |
| 6 | Austria | 647111 | 334629 | 361 | 1646 | 7697 | 5295 | 2272 | 1596 | 0 |
| 7 | Azerbaijan | 336788 | 199127 | 11 | 34 | 147 | 543 | 276 | 169 | 7 |
| 8 | Bahrain | 266797 | 90062 | 195 | 256 | 473 | 9 | 1 | 5 | 1 |
| 9 | Bangladesh | 1000543 | 499560 | 3 | 10 | 48 | 15144 | 11758 | 2184 | 7 |
| 10 | Belarus | 424554 | 171579 | 12 | 46 | 94 | 3786 | 954 | 91 |  |
| 11 | Belgium | 1092477 | 625928 | 314 | 1486 | 7284 | 6637 | 2458 | 478 | 0 |
| 12 | Bhutan | 2258 | 446 | 1 | 1 | 3 | 4 | 3 | 0 | 0 |
| 13 | Bolivia | 449687 | 149149 | 3 | 12 | 61 | 14758 | 3364 | 2619 | 120 |
| 14 | Brazil | 18909037 | 7162978 | 77 | 291 | 2915 | 377176 | 59124 | 60607 |  |
| 15 | Brunei Darussalam | 266 | 152 | 12 | 56 | 115 | 21 | 2 | 1 | 0 |
| 16 | Bulgaria | 422353 | 191029 | 7 | 92 | 293 | 5067 | 1817 | 601 | 0 |
| 17 | Burkina Faso | 13505 | 4954 | 2 | 26 | 146 | 843 | 0 | 186 | 0 |
| 18 | Cameroon | 80858 | 25849 | 2 | 10 | 75 | 1373 | 705 | 243 | 1 |
| 19 | Canada | 1418632 | 495346 | 138 | 569 | 4018 | 90839 | 9066 | 5116 |  |
| 20 | Chile | 1579591 | 583354 | 33 | 238 | 1610 | 9831 | 4424 | 1436 | 0 |
| 21 | China | 119141 | 95716 | 80991 | 81174 | 82230 | 1773689 |  |  |  |
| 22 | Colombia | 4426811 | 1482072 | 9 | 93 | 491 | 37773 | 7214 | 5188 | 0 |
| 23 | Costa Rica | 377091 | 157472 | 22 | 50 | 231 | 12811 | 5871 | 2101 | 0 |
| 24 | Cote d Ivoire | 48693 | 21772 | 1 | 9 | 92 | 93 | 8 | 75 | 0 |
| 25 | Croatia | 360680 | 194962 | 16 | 81 | 586 | 1676 | 752 | 127 | 0 |
| 26 | Cyprus | 81581 | 17476 | 6 | 58 | 162 | 1141 | 824 | 155 | 0 |
| 27 | Czechia | 1668891 | 624140 | 116 | 522 | 2279 | 4092 | 1319 | 236 | 4 |
| 28 | Denmark | 297543 | 131606 | 674 | 1044 | 2046 | 2629 | 438 | 46 | 0 |
| 29 | Dominican Republic | 331826 | 159064 | 5 | 21 | 581 | 7070 | 2525 | 972 | 80 |
| 30 | Ecuador | 465878 | 205920 | 17 | 155 | 1595 | 60733 | 21329 | 10346 | 0 |
| 31 | Egypt | 282582 | 124891 | 67 | 196 | 536 | 8044 | 3599 | 3199 | 0 |
| 32 | Estonia | 131396 | 21794 | 13 | 258 | 575 | 636 | 107 | 29 |  |
| 33 | Finland | 97651 | 32582 | 109 | 359 | 1025 | 1073 | 0 | 22 |  |
| 34 | France | 5686066 | 2418439 | 2860 | 9043 | 32542 | 85072 | 39112 | 6180 | 0 |
| 35 | French Polynesia | 19026 | 16182 | 1 | 3 | 30 | 27 | 3 | 6 | 0 |
| 36 | Georgia | 374836 | 208638 | 25 | 38 | 85 | 2237 | 1516 | 305 | 20 |
| 37 | Germany | 3734468 | 1494009 | 2369 | 8198 | 48582 | 44948 | 11687 | 16125 | 5 |
| 38 | Greece | 433021 | 130485 | 98 | 418 | 966 | 9932 | 2014 | 2258 | 1327 |
| 39 | Guyana | 20645 | 6076 | 1 | 4 | 8 | 359 | 13 | 52 | 0 |
| 40 | Honduras | 270689 | 116212 | 2 | 9 | 67 | 7195 | 758 | 102 | 0 |
| 41 | Hungary | 808437 | 302989 | 16 | 58 | 343 | 8539 | 3535 | 793 | 0 |
| 42 | Iceland | 6675 | 5621 | 61 | 250 | 890 | 2 | 0 | 0 | 0 |
| 43 | India | 30752950 | 10031223 | 74 | 151 | 724 | 58160 | 13055 | 20619 | 0 |
| 44 | Indonesia | 2455912 | 657948 | 34 | 227 | 1046 | 1597 | 224 | 929 | 53 |
| 45 | Iran | 3327526 | 1152072 | 10075 | 17361 | 32332 | 6841 | 1100 | 1756 | 5 |
| 46 | Iraq | 1406289 | 583118 | 70 | 164 | 458 | 215 | 12 | 85 | 3 |
| 47 | Ireland | 276104 | 78776 | 70 | 292 | 2121 | 2625 | 602 | 29 | 0 |
| 48 | Israel | 845123 | 371373 | 75 | 427 | 3460 | 6093 | 2843 | 525 | 0 |
| 49 | Italy | 4267105 | 1938083 | 15113 | 35713 | 86498 | 54153 | 31398 | 10464 | 454 |
| 50 | Jamaica | 50497 | 12135 | 1 | 13 | 26 | 621 | 177 | 204 | 15 |
| 51 | Japan | 813976 | 195880 | 675 | 873 | 1499 | 52332 | 21461 | 17125 | 3 |
| 52 | Jordan | 754927 | 272797 | 1 | 52 | 235 | 675 | 216 | 391 | 0 |
| 53 | Kuwait | 369227 | 147775 | 80 | 142 | 235 | 32 | 4 | 14 | 12 |
| 54 | Latvia | 137797 | 30297 | 16 | 71 | 280 | 1568 | 213 | 36 | 0 |
| 55 | Lebanon | 546366 | 156570 | 66 | 133 | 391 | 1816 | 988 | 255 | 39 |
| 56 | Lithuania | 279024 | 112359 | 3 | 26 | 358 | 2049 | 677 | 57 | 0 |
| 57 | Luxembourg | 71879 | 44067 | 17 | 210 | 1605 | 148 | 76 | 24 | 0 |
| 58 | Malaysia | 808658 | 91969 | 129 | 673 | 2161 | 44115 | 3021 | 3547 | 94 |
| 59 | Maldives | 74848 | 13474 | 8 | 13 | 13 | 363 | 51 | 279 | 30 |
| 60 | Malta | 30851 | 11621 | 9 | 48 | 139 | 89 | 83 | 3 | 0 |
| 61 | Mexico | 2558369 | 1301546 | 12 | 93 | 589 | 53144 | 28601 | 12991 | 7 |
| 62 | Mongolia | 131962 | 953 | 1 | 5 | 12 | 116 | 10 | 5 |  |
| 63 | Morocco | 537253 | 415226 | 6 | 49 | 358 | 13697 | 3965 | 7775 | 295 |
| 64 | Nepal | 652859 | 253184 | 1 | 1 | 3 | 574 | 247 | 181 | 11 |
| 65 | Netherlands | 1701911 | 676589 | 614 | 2051 | 8603 | 9309 | 4288 | 254 | 0 |
| 66 | New Zealand | 2409 | 1760 | 5 | 20 | 416 | 5086 | 1296 | 303 | 0 |
| 67 | North Macedonia | 155760 | 77949 | 7 | 36 | 219 | 98 | 60 | 27 |  |
| 68 | Norway | 132572 | 42775 | 489 | 1423 | 3581 | 614 | 83 | 13 | 22 |
| 69 | Oman | 281688 | 127019 | 18 | 33 | 152 | 204 | 29 | 112 | 0 |
| 70 | Pakistan | 967633 | 454673 | 20 | 241 | 1235 | 1 | 0 | 0 |  |
| 71 | Panama | 411226 | 206310 | 14 | 86 | 674 | 2403 | 551 | 160 | 0 |
| 72 | Paraguay | 432801 | 98296 | 5 | 11 | 52 | 19662 | 4539 | 2834 | 0 |
| 73 | Peru | 2071637 | 993760 | 22 | 145 | 580 | 4800 | 1189 | 705 | 15 |
| 74 | Philippines | 1455585 | 458044 | 52 | 187 | 803 | 12595 | 4463 | 1100 | 14 |
| 75 | Poland | 2880670 | 1202700 | 49 | 287 | 1389 | 22742 | 7992 | 1770 | 0 |
| 76 | Portugal | 899295 | 370787 | 41 | 642 | 4268 | 8172 | 4183 | 2084 | 0 |
| 77 | Qatar | 222918 | 141858 | 262 | 442 | 562 | 68 | 8 | 60 |  |
| 78 | Republic of Korea | 165344 | 49665 | 7979 | 8413 | 9478 | 18716 | 5576 | 5674 |  |
| 79 | Republic of Moldova | 257272 | 134578 | 4 | 36 | 199 | 2853 | 1317 | 391 | 1 |
| 80 | Réunion | 31845 | 8704 | 3 | 12 | 135 | 5141 | 1760 | 641 |  |
| 81 | Russian Federation | 5733218 | 2848377 | 34 | 147 | 1264 | 76369 | 26164 | 10198 | 253 |
| 82 | Saudi Arabia | 497773 | 360848 | 21 | 238 | 1104 | 5413 | 2011 | 3131 | 0 |
| 83 | Senegal | 44790 | 17670 | 10 | 36 | 119 | 479 | 20 | 85 | 2 |
| 84 | Slovakia | 391813 | 151336 | 21 | 105 | 295 | 1786 | 469 | 137 | 1 |
| 85 | Slovenia | 257619 | 105013 | 57 | 286 | 632 | 1172 | 849 | 55 | 0 |
| 86 | South Africa | 2135246 | 912477 | 17 | 116 | 1170 | 26857 | 8928 | 6158 |  |
| 87 | Spain | 3915313 | 1797236 | 2965 | 13716 | 64059 | 61343 | 38067 | 6488 |  |
| 88 | Sri Lanka | 269946 | 36049 | 3 | 42 | 106 | 2260 | 818 | 255 | 0 |
| 89 | Sweden | 1092308 | 367120 | 620 | 1279 | 3046 | 1459 | 209 | 46 |  |
| 90 | Switzerland | 701473 | 402264 | 858 | 3010 | 12104 | 2077 | 1012 | 303 | 0 |
| 91 | Thailand | 317506 | 4331 | 75 | 212 | 1136 | 35287 | 0 | 21601 | 0 |
| 92 | Togo | 14176 | 3396 | 1 | 1 | 25 | 1293 | 19 | 522 | 7 |
| 93 | Tunisia | 464914 | 119151 | 7 | 29 | 227 | 1003 | 645 | 236 | 9 |
| 94 | Turkey | 5465094 | 1189947 | 1 | 191 | 5698 | 60020 | 23047 | 16069 | 309 |
| 95 | Ukraine | 2240246 | 964448 | 3 | 16 | 311 | 25341 | 4802 | 1806 | 13 |
| 96 | United Kingdom | 5022897 | 2004223 | 594 | 2630 | 14547 | 19301 | 6506 | 376 | 0 |
| 97 | United States of America | 33451965 | 17314834 | 1264 | 7087 | 85228 | 407779 | 24040 | 65771 | 0 |
| 98 | Vietnam | 24810 | 1411 | 39 | 66 | 169 | 19154 | 5391 | 9661 | 8 |
| Correlation | Total Pesticides | 0.21 | 0.23 | 0.93 | 0.86 | 0.57 |  |  |  |  |
|  | Total Fungicides and Bactericides | 0.54 | 0.52 | 0.30 | 0.42 | 0.51 |  |  |  |  |
|  | Total Insecticides | 0.80 | 0.83 | 0.11 | 0.18 | 0.47 |  |  |  |  |
|  | Total Rodenticides | 0.00 | 0.00 | 0.21 | 0.22 | 0.14 |  |  |  |  |

Pests, pathogens, and weeds pose a major challenge to agricultural production, and pesticides are widely used to ensure food security across the world. However, the toxicological effects of pesticides on the human health is subjected to many reviews via multiple pathways involving food, drinking water, residential, occupational [19-21]. At this moment COVID-19 outbreak is resulting more than 250 plus global fatality every hour in human affairs and its counter response is weak in many countries. Outcome from our unbiased scientific data analysis reveal the use of pesticides has appeared as the third most contributor (10.80%) in case of COVID-19 incidences.

A report by the Food and Agriculture Organization of the United Nations (FAO) suggests that China is the world’s largest consumer of agricultural chemicals and country, which alone uses more than 30% of global fertilizers/ pesticides in 9% of global cropland to fulfill the high food demands. Second pesticides consumer United States, and third, Brazil, today as on 09-July-2021 are with first-three largest COVID-19 infected cases 33451965, and 18909037 respectively. Further 4th pesticide consumer country, Argentina supports for a high 4593763 cases, in our opinion country Argentina is at a high COVID-19 risk during the writing of this paper. Another leading pesticide consumers Canada(5th), Ukraine(6th), France (7th) etc. are currently with high outbreaks of COVID-19. On the other hand, wholly organic Bhutan reports just 2258 COVID-19 infected individuals and most of them are imported cases only. In India, the largest pesticide consumer state Maharashtra as of July 11th, 2021 is with the highest 117270 total active COVID-19 cases, whereas wholly organic Indian state Sikkim is with a low number of 2244 cases. Countries with high population density including Pakistan, Bangladesh, Nigeria, etc. are not that much affected as leading pesticide consumers, provides further evidence to redefine multifactorial approach in COVID-19. A handful of healthy soil comprised of millions of ecological communities of microbes (microbiota), also regarded as a protective shield against pathogen invasion.

Pesticides are non-specific=> inevitably use kills both pests as well as protective microbiota in the surrounding soil, and water sources => a loss in biodiversity (Fig. 1). Thus, a lower amount of pesticide consumption-led drop in COVID-19 cases and vice-versa. Nonetheless, pesticides sold in Africa is less than 4% of global pesticide trade and African continent is noticeable with low COVID-19 infected cases all over. Overall, a scientific dose-response relationship is fitting well for pesticide consumption and active COVID-19 cases. Therefore, it could be easily postulated that the generous use of pesticides might be responsible to trigger the high transmissibility of the SARS-CoV-2 virus.

**Figure 1.**
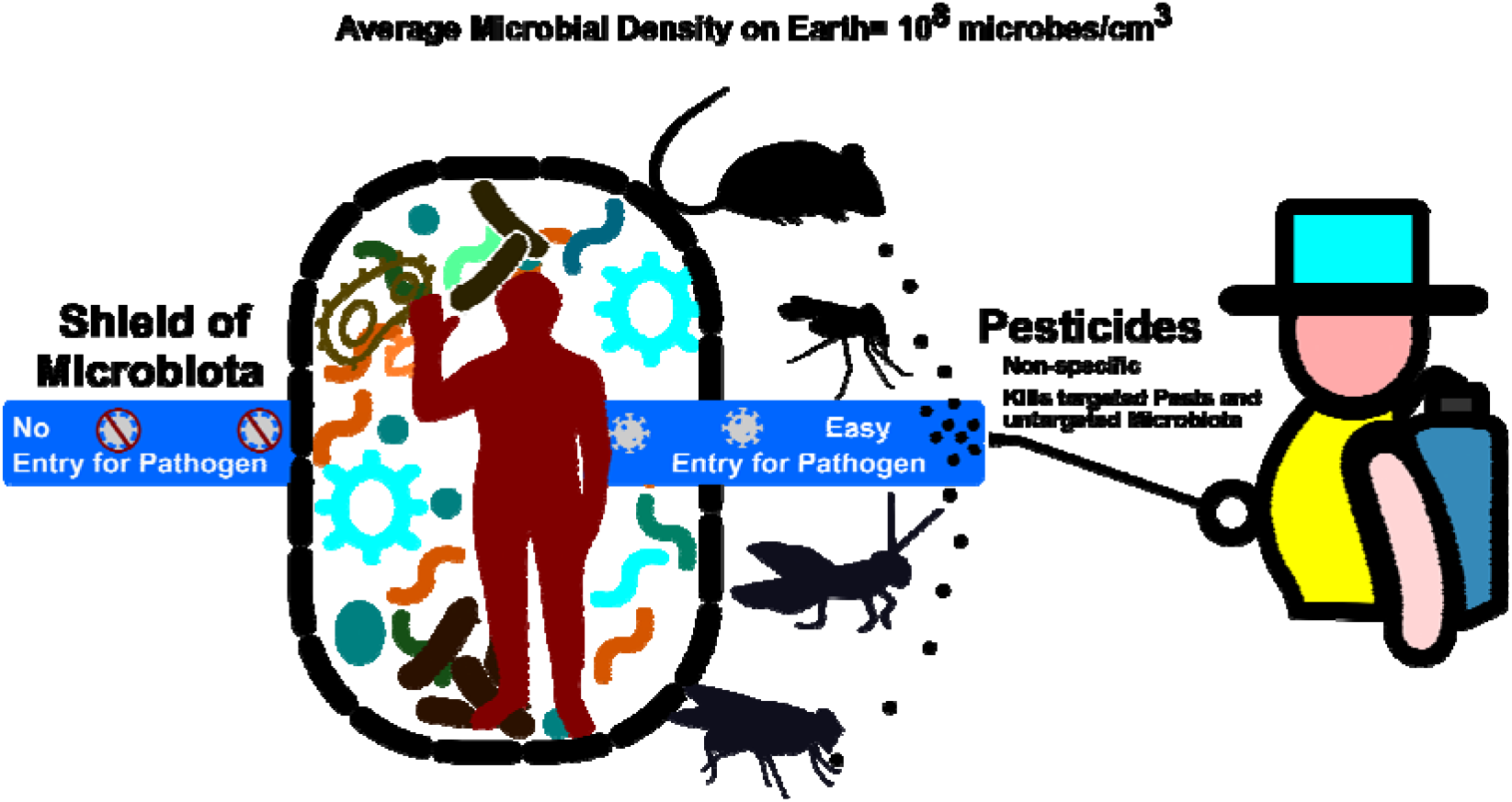
Mechanism of Pesticide Use over COVID-19 progression

In fact, two researchers, Lusk and Chandra highlighted during the beginning of COVID-19 (from March 1, 2020, to March 31, 2021), a reduction in agricultural output by about $309 million in the United States, all due to a reduction in labor availability[22]. The study also found that the average incidence and death incidence rate in farmers/farmworkers amounted to 9.32 % and 0.17% respectively. Both average incidence and death incidence rates were raised as high as about 3-fold in farmers/ farm workers in comparison to other normal individuals. Unfortunately, the cause of high COVID-19 outbreaks in farmers/farmworkers remains unpredictive. This evidence clearly suggests that farmers/ farm workers working near pesticide agricultural places are potentially at a very high risk for contracting COVID-19.

The use of household insecticides in form of spray-cans, oil-sprays, mosquito coils and vaporizing systems are widely used to kill cockroaches, mosquitoes, bedbugs, beetles, ants etc. With increased urbanization and awareness of home pest control methods all over, for instance, New York US, Bergamo Italy, Community of Madrid Spain, New Delhi India, Sao Paulo Brazil etc. have become favorable for insecticide consumption. These places have already become epicenter of COVID-19 outbreaks.

The SARS-COV2 virus is continually mutating and this natural evolution led increased transmissibility, virulence, and resistance to antibody neutralization poses. Recently as on 14-Jun-2021, eight variants of interest *viz*. α, β, γ, δ, η, ι, κ, and λ are listed by the WHO successfully [23]. SARS-CoV-2 virus contains the genetic element s2m and as reported by the Tengs andcoworkers, recently xenologue of s2m was found in a large number of insect species [24]. As a result, the earlier record of a total pesticides is recently shifted to insecticide use in this report.

Undoubtedly, microbiota represents the first line of defense and protection against pathogens. For instance, presence of commensal microbiome in human skin offers temperature regulation, ultraviolet (UV) radiation protection, vitamin D production, and most importantly, keeping pathogenic microbes outside the body. The pesticides are non-specific and its indiscriminate use leads to the degradation of useful microbiota. Which may likely to increase the harmful pathogen entry in human. Recent finding with high correlation values are in full agreement that local insecticide consumer sites may likely to increase the high transmissibility of the SARS-CoV-2 virus.

Formal education and counseling of farmers and other stake-holders on pesticide use are urgently needed. Conscious monitoring of pesticide use worldwide (For harmonious coexistence of micro-/macro-organisms in nature). In line with modern science multi-factorial approach, this study is endorsing nature’s role in direct pathogen invasion and to limit the transmission of SARS-COV-2 pathogen. Thus, similar approach help researchers think undoubtedly about the causation of COVID-19 disease. We hope these insights presented above along with disease reduction framework will be a game-changer in fighting against the COVID-19 pandemic.

## Data Availability

WHO Situation report - 148 Coronavirus disease 2019 (COVID-19) 
Food and Agriculture Organization (FAO) report  

https://www.who.int/docs/default-source/coronaviruse/situation-reports/20200616-covid-19-sitrep-148-draft.pdf?sfvrsn=9b2015e9_2

http://www.fao.org/faostat/en/#data/RP

## Acknowledgements

The authors are thankful to the Food and Agriculture Organization (FAO) for keeping data free to use. The authors are also thankful to the GEHU India for providing necessary facilities and technical support.

## Author contributions

AA and RR collected and verified data. PG designed and GJ analyzed the data. PG drafted the manuscript. All authors critically revised the manuscript and approved the final version for submission.

